# The Lifecycle of Electronic Health Record Data in HIV-Related Big Data Studies: A Qualitative Study of Instances of and Potential Opportunities to Minimize Bias

**DOI:** 10.1101/2024.09.30.24314636

**Authors:** Arielle N’Diaye, Shan Qiao, Camryn Garrett, George Khushf, Jiajia Zhang, Xiaoming Li, Bankole Olatosi

**Author notes:** **Corresponding Author:** Shan Qiao.

## Abstract

**Background:** Electronic health record (EHR) data are widely used in public health research, including HIV-related studies, but are limited by potential bias due to incomplete and inaccurate information, lack of generalizability, and lack of representativeness. This study explores how workflow processes within HIV clinics, among data scientists, and within state health departments may introduce and minimize bias within EHRs.

**Methods:** Using a constructivist grounded theory approach, in-depth individual interviews were conducted with 16 participants purposively sampled in South Carolina from August 2023-April 2024. A focus group with 3 health department professionals with expertise in HIV disease surveillance was also conducted. Analysis was conducted as outlined by Charmaz (2006).

**Results:** To reduce bias in EHR data, information entry forms should be designed to expansively include patient self-reported social determinants of health (SDOH) information. During data collection, healthcare providers should create a supportive healthcare environment, facilitate SDOH information disclosure, and accurately document patient information. Patients should have access to their EHRs to confirm that SDOH information are correctly recorded. During data curation, data scientists should inspect datasets for completeness, accuracy, and educate public health researchers on dataset limitations. During data management and utilization, health department professionals should crossmatch data across the state, customize data collection systems to reflect local needs, and provide community-based data education and stigma management.

**Conclusion:** Study results suggest that future research is needed to understand how healthcare systems can be incentivized to create and implement EHR bias reduction strategies across all workflows and between stakeholders.

## Introduction

Electronic health record (EHR) data is widely used in public health research, including HIV-related studies. In HIV research, EHR data is used for disease surveillance, to understand treatment uptake, to assess the efficacy of treatment regimens, and to examine health outcome disparities (1-3). As a data source, EHR data is viewed as an attractive option because of its cost effectiveness, when compared to primary data collection, the high volume of population level data it yields, and its suitability for multidimensional analyses (1, 2, 4). However, existing literature notes that EHR, as a data source, is limited by potential bias from incomplete and inaccurate information, lack of generalizability, and underrepresentation (4). Many scholars attribute this to EHR being designed for medical billing, scheduling, and clinical record keeping rather than for public health research (4). Scholars also attribute EHR data’s limitations to its vulnerability to social biases, influencing both how and if information is entered into EHR systems (5, 6).

Current literature notes that EHR data used for HIV research is vulnerable to biases during data collection given most clients of HIV-related care services endure stigma, discrimination, and other adversities in relation to the social determinants of health (3). In healthcare settings (i.e., both HIV-specific and non-HIV-specific), these biases are observed as stemming from institutional policies, training practices, and healthcare provider biases—in-turn influencing the accuracy, completeness, and representativeness of EHR data (5, 6). Within HIV prevention, the implicit biases of healthcare providers, via their interpretation and EHR system documentation of patients’ sexual history, exacerbated patient challenges with accessing HIV prevention resources such as pre-exposure prophylaxis and HIV counseling (7-9). Scholars observe this as disproportionately affecting lesbian, gay, bisexual, transgender, and queer (LGBTQ) individuals, women, and racial minorities (8, 9). The current literature offers a variety of techniques to address bias derived from inaccurate, incomplete, and under-representative data. These techniques include imputation of surrogate measures to address missing information, conducting validation studies with external datasets to determine the representativeness of EHR data, and performing sensitivity analyses to identify misclassified information. Additional techniques include using causal diagrams to support causal inferences and using record audits to assess data quality (10-13).

Beyond employing advanced data science analytics, recent literature also highlights the importance of understanding how workflows are structured to identify and minimize opportunities for EHR data bias across various stakeholder perspectives (e.g., data scientists, public health professionals, healthcare providers, and patients). Richesson (14) found that 10 out of the 20 clinical trial sites, featured within their study on EHR research settings, lacked standardized workflows for the extraction, preparation, and use of EHR data for research, subsequently warranting further research on best practices (14). In addition, there is a lack of studies regarding the lived experiences of diverse key stakeholders who are engaged in different stages of the EHR data lifecycle, including HIV patients, healthcare providers, and data scientists (15). This study aims to fill the above-mentioned knowledge gaps, by demonstrating the workflows of EHR data between patients, healthcare providers, and data scientists as well as by exploring where bias is introduced within the workflow processes between HIV clinics, public health departments, and among data scientists and where opportunities may exist to minimize this bias within EHR data collection and utilization. This study had the overarching research question of: *How is bias introduced within the workflows of healthcare and public health research settings and how might they be structured to minimize opportunities for bias in EHR big data?*

## Materials and Methods

### Theoretical Approach and Research Paradigm

This study employed a constructivist grounded theory approach as outlined by Charmaz (16). As an approach, constructivist grounded theory holds the worldview that researchers actively play a role in defining and shaping meanings derived from the data, as created by study participants during their interviews and focus groups (17). This theoretical approach was also chosen because it was best suited to answer this study’s research question.

### Reflexivity: Researcher Characteristics

All research team members have formal training in qualitative research methods and have previously conducted HIV and social determinants of health specific studies. Additionally, all research team members conduct research in an academic setting and have either worked with or have been exposed to research that uses EHR data. Five authors (SQ, KH, BO, ZJ, XL) have doctorate degrees (i.e., PhD) and two authors (AN, CG) are doctoral students who have completed an undergraduate degree in sociology (AN, CG) and public health (CG) (i.e., BA) as well as a master’s degree in public health (i.e., MSPH, MPH).

### Sampling Strategy

Participants were purposively sampled to represent the perspectives of patients living with HIV, healthcare providers, data scientists who provide EHR datasets to public health researchers, and health department professionals who work with EHR datasets. Patients living with HIV had the inclusion criteria of being at least 18 years of age and living with HIV in South Carolina. Healthcare providers had the inclusion criteria of being at least 18 years of age and working within HIV-specific facilities (e.g., HIV clinics) or non-HIV healthcare facilities in South Carolina that use EHR systems. Data scientists had the inclusion criteria of being at least 18 years of age and being a professional data curator, data management expert, or data repository administrator. Public health department professionals working in HIV surveillance and retention in care had the inclusion criteria of being at least 18 years of age, working in state agencies overseeing and administering public data repositories in South Carolina, and being engaged in EHR data management and utilization.

All participants were purposively recruited through referrals from a local research partner network, which consists of stakeholders from AIDS service organizations, community-based organizations, academia, state public health agencies, people living with HIV, and state policy makers. After contacting a member of the research team, participants were provided with an invitation outlining study activities and the voluntary nature of participating in the present study. This recruitment strategy yielded 16 participants (i.e., 5 patients, 5 healthcare providers, 5 data scientists, and 1 health department professional providing retention-in care services). This recruitment strategy also yielded a focus group (n=1) with 3 health department professionals working in HIV disease surveillance. Study participants were recruited from August 2023-April 2024 and received a $50 gift card for their participation in this study.

### Data Collection

Individual in-depth interviews (n=16) were conducted by research team members using a semi-structured interview guide. Six interviews were conducted over video conference (e.g., Zoom, Microsoft Teams) and ten interviews were conducted in-person. Interviews were conducted using both mediums because existing literature finds both virtual and in-person interview formats to be comparable (18). Virtually interviewed participants were advised to be in a location where they were comfortable and able to have a private conversation. In-person interviews were conducted in a private room at a location that was convenient for the participant. Interviews ranged from 45 to 60 minutes in length and were audio recorded. The focus group (n=1) was conducted in-person in a private office at the participants’ workplace. The focus group lasted for 73 minutes and used the same semi-structured interview guide as the individual in-depth interviews.

### Data Analysis

Transcripts were first transcribed verbatim using Otter.ai (version 3.43.2-240212-89103881). Transcripts were then checked by members of the research team for accuracy and de-identified of all identifying information. Individual interviews were the unit of analysis.

Transcripts were first initially coded, where sentences and paragraphs were labeled with codes. These codes were then focus, axial, and theoretically coded (16). Throughout the analytical process, constant comparison was used to identify emergent trends within the data. Analysis was first conducted by one member of the research team (AN). Codes and latent level categorizations were then reviewed and discussed with author (CG). An analytical matrix of study findings was used to establish consensus among all authors (AN, SQ, CG, KH, BO, ZJ, XL). A reflexive journal was kept by AN throughout the data analysis process of this study. This journal was discussed when interpreting study results to prevent the introduction of potential researcher biases. An audit trail was used throughout data collection and analysis to increase the credibility of study results. This study was reported using the Standards for Reporting Qualitative Research (SRQR) Checklist (19).

### Ethical Considerations

This study received IRB approval from The University of South Carolina Office of Research Compliance (Application No.: Pro00122501) and was conducted in accordance with the Declaration of Helsinki (20). All participant audio recordings and original transcripts were stored in a dual authentication, password protected cloud drive, and made accessible only to members of the research team. At the beginning of each interview, participants received the invitation to participate and gave verbal consent; they were not required to give written consent. Furthermore, at the start of each interview, participants were assigned a participant ID that corresponded with their respective transcripts to ensure their anonymity.

## Results

The findings of the present study describe instances where bias is introduced over the EHR data lifecycle, ways in which stakeholders work to mitigate biases, and their recommendations for structural interventions to mitigate bias (Figure 1). In clinical settings (i.e., data collection), healthcare providers describe the influence of socio-structural biases on their inquiry, interpretation, and documentation of patients’ social determinants of health (SDOH) information. Data scientists, within their workflow (i.e., data curation), illustrate the implications of limited data availability and representativeness in biasing the data they manage. Health department professionals, within their workflow (i.e., data management and utilization), describe the challenges of using delayed and incomplete data to create research products and public health decision making tools. It is important to note that participants described limitations to reducing bias as their workflows overall were 1) not intentionally structured to minimize opportunities for bias and 2) because EHR data is touched by many different stakeholders both within and between workflows. These touch points are summarized in Figure 2.

**Figure 1.**
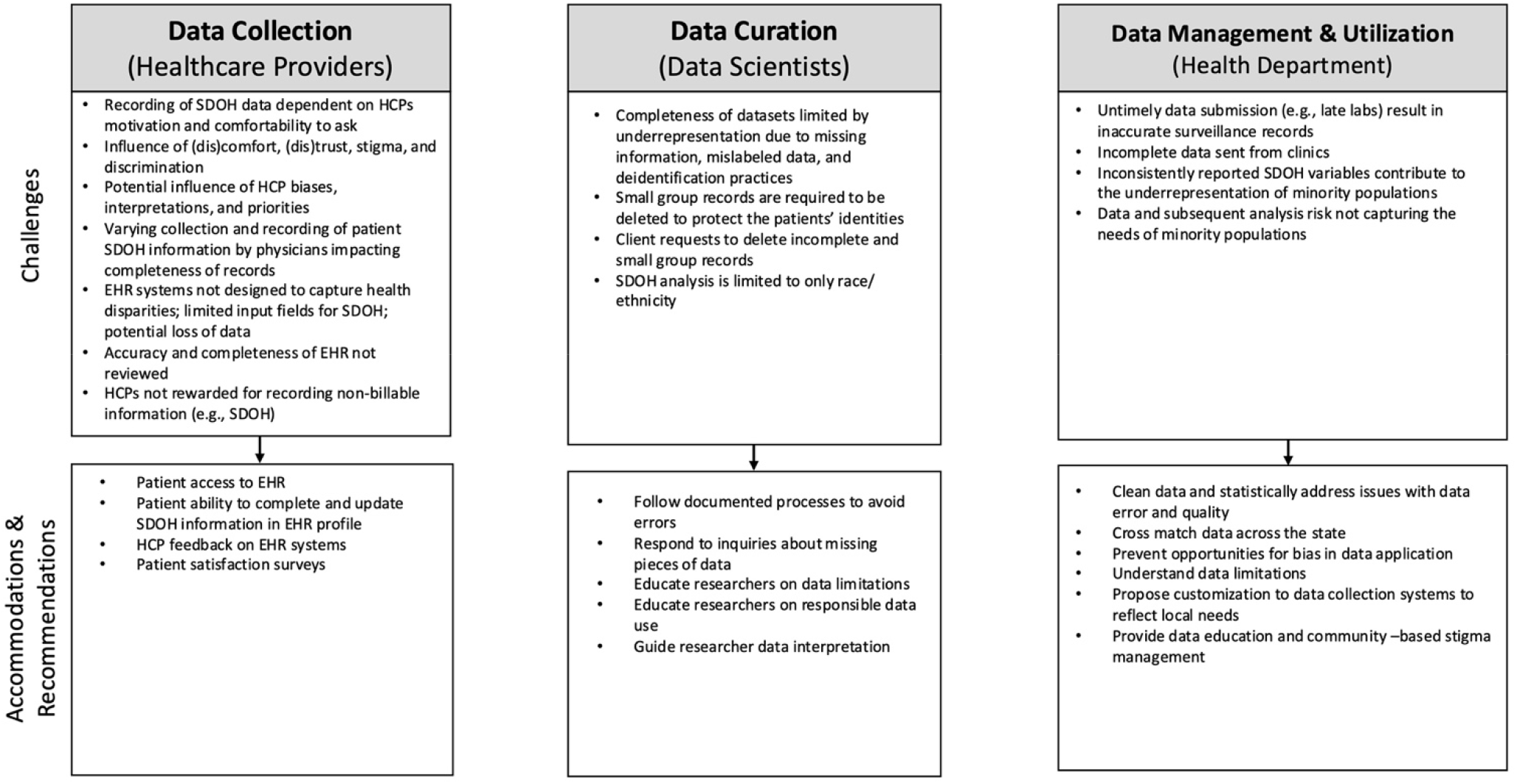
Challenges, accommodations, and recommendations for EHR data bias reduction.

**Figure 2.**
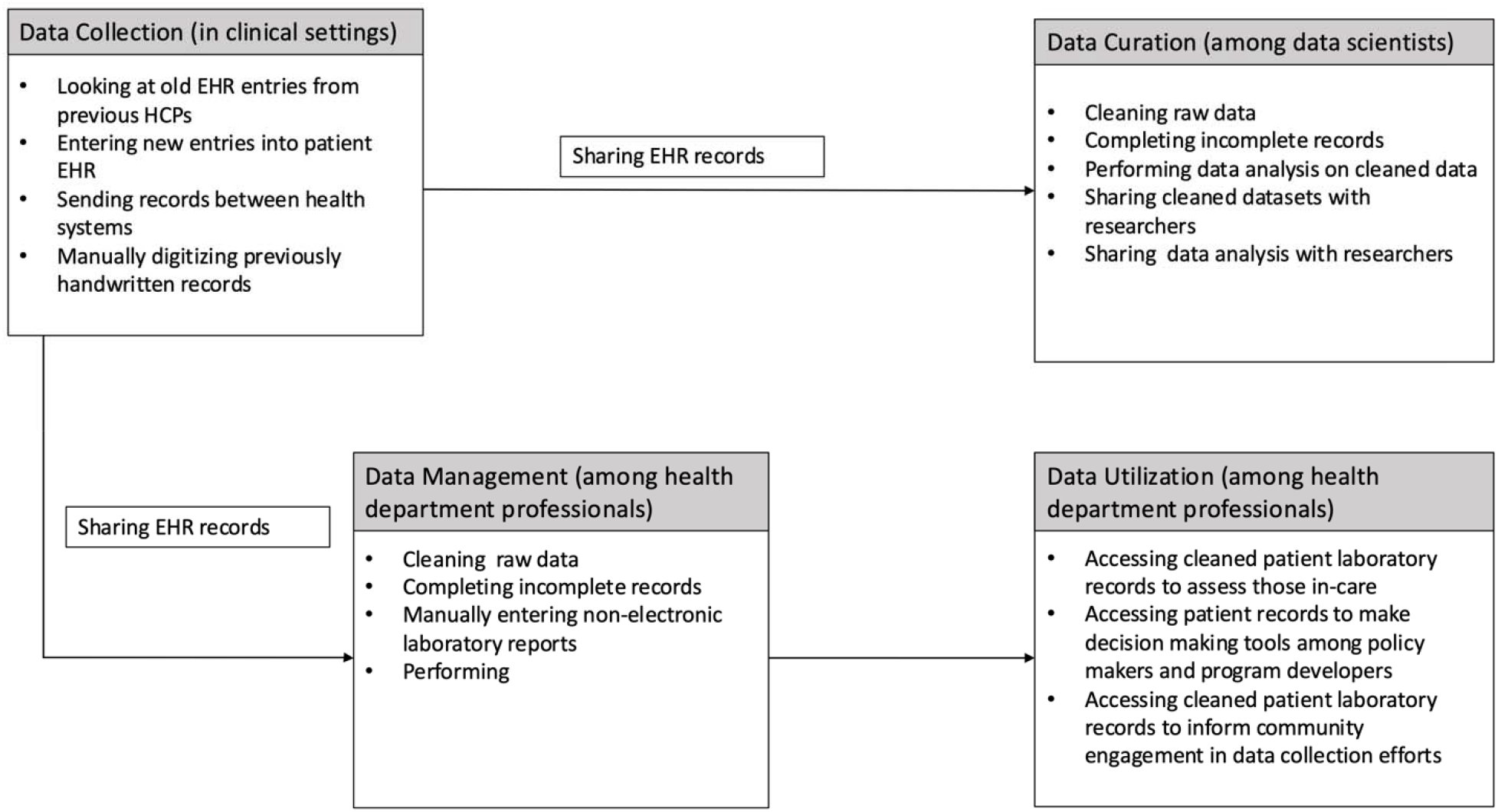
Description of data touchpoints throughout the EHR data lifecycle.

### Data Collection: In Clinical Settings During and After Patient Appointments

During data collection at patient medical appointments, the recording of patient SDOH information was dependent on a variety of factors: health provider motivation and comfortability to ask patients about their SDOH information as well as patient comfort and trust with their healthcare provider. Both healthcare provider and patient participants described HIV clinical settings as being conducive to making both patients and providers feel comfortable discussing patient specific information. Healthcare provider 1 describes:

> *“Again, I can’t speak to other offices or other clinics who do the same thing that we do [at the HIV clinic]. But a lot of times with our providers, they’re most often extremely sensitive to the social thing. And so oftentimes, we run behind on time, right? Because providers have a certain amount of time that they are allotted to spend with a patient. And it’s almost never that short. It’s always grows beyond that, and that a lot of times is hinged on the social stuff on these conversations that the patients will have that a little bit beyond just you taking your medication? Cool. How do you feel here? Let me check this. Let’s get labs, right*.*”* (Healthcare provider 1)

This is contrast to non-HIV clinical settings where both healthcare provider and patient participants described these settings as promoting patient and provider discomfort when discussing patient SDOH information, promoting patients to feel stigmatized by healthcare providers, and disincentivizing providers from both inquiring about and documenting patient SDOH information. Among providers, this dis-incentivization was attributed to some healthcare systems not rewarding the recording of non-billable information. This was also attributed to many EHR systems not having a standard field to enter information that is inclusive of sexual and gender minorities (e.g., transgender, non-binary, preferred/lived name vs. dead name). In such settings, healthcare providers described experiencing variability in what, when, and how their colleagues documented patient SDOH information. Whereby, some healthcare providers did not record patient SDOH information, others recorded some SDOH information, and others recorded only what they perceived as being relevant. Healthcare provider 2 and Patient 1 note:

> *“But, you know, like, I don’t put in anyone’s address. I don’t put anyone’s phone number. I don’t put anyone’s ethnicity. I don’t put in anyone’s gender. I don’t, I know that you can put in that they’re transgender, you could put in their preferred name. Sometimes they’ll have a full name change. Sometimes it’ll be put in as a nickname. Sometimes you’ll have their dead name and then their preferred name as in quotations after too, so it depends on how it’s done and who does it*.*”* (Healthcare provider 2)
>
> *“So, when you meet an older physician, if they get surprised when they finally look at you and they see like oh, okay, okay, well, this is what you are assigned at birth. So this is how I’m going to speak to you. It doesn’t matter. [Good]-bye your pronouns, this is what you’re assigned at birth, so this is what I’m gonna refer to you as. I don’t know how to deal with that, I don’t know how to speak to you. And this is just what I’m used to, this is how I was trained, so this is what it’s going to be*.*” (Patient 1)*

Within the HIV clinical setting, the checking of charts by healthcare providers was a facilitator for reducing opportunities for bias. These charts were reviewed by providers to check the notes they had written, to review the work of others for compliance, or to solely review records for further information. Healthcare provider 3 describes:

> *“[…] I check the schedule every day [and] I go through and look at who needs to meet with the case manager and I assigned case managers. I’ll go and look for, coverage, people who don’t have insurance, people who have Medicaid, and people who are connected to our drug assistance program, who maybe need to update their file [regarding] intakes, assessments, treatment plans and things like that to make sure that we’re compliant with our program. And so I go through the charts that way in [name of a type of EHR system], and it will say transgender male to female or vice versa, So it’s recorded*.*”* (Healthcare provider 3)

According to healthcare provider participants, this is in contrast to their experiences outside of the HIV clinical setting, where they reported records not being checked and varying in quality. Within non-HIV clinical settings, this was attributed to not having the quality control of patient records fall within the purview of anyone’s job. Healthcare provider 4 observes:

*“Should is a funny word. I think that there should be committed individuals who do clinical care, who have a portion of their time paid for to commit to this work. And I think that departments should have a champion for this kind of work. And working and navigating the medical record in general. Just like in our division, we have champions for antibiotic stewardship, and infection prevention about like spreading diseases around the hospital, and all these other things, you know, we wear those hats, those are recognized as part of the work that we do administratively for the hospital*.*”* (Healthcare provider 4)

When asked about recommendations for minimizing opportunities for EHR bias within clinical settings, both patient and healthcare provider participants recommended that patients have access to their EHR records, through a patient portal, where patients can update identity specific information (i.e., preferred name, gender identity, sexual orientation). Additionally, patient participants recommended care experience surveys be used to help capture patient experiences of bias during their medical appointments. Furthermore, healthcare provider participants discussed how they should have an opportunity to provide feedback on their experience using EHR systems. Among these participants, most of them expressed having never been asked for their feedback on EHR systems before. This is captured by healthcare provider 1 who, when asked if they’ve ever been asked for feedback about what should be recorded in EHR systems and the best methods for conveying this information, replied *“No one’s asked me”* (Healthcare provider 1).

### Data Curation by Data Scientists

In their workflows, data scientists attempted to address opportunities for statistical biases in the raw data they received from hospital systems (i.e., both HIV-specific and non-HIV-specific). Following the reception of raw EHR data from medical systems, data scientist participants described first encrypting and assigning de-identifying labels to patient data. Data scientists then described a process for cleaning the data, including checking it for general issues and missing values. According to data scientist participants, this data cleaning process followed a documented protocol in order to avoid opportunities for error. Data scientists 1 and 2 observe:

> *“How we avoid errors and that we do have a document that describes our process, generally, of how that that unique identification number is assigned, that uses a combination of data elements, such as first name, last name, social security number and date of birth, but it also can use things like race and sex*.*”* (Data scientist 1)
>
> *“Yeah, with the healthcare stuff, it it’s a lot of like cleaning up data, organizing it. Putting reports together for the hospitals or for [name of a hospital association][…]”* (Data scientist 2)

Once completed, data scientist participants reported their bias minimization process as shifting to focus on data application.

After receiving data access requests from researchers, data scientists’ workflows centered around minimizing opportunities alongside the recipient. According to these participants, this occurred through creating and publishing a data dictionary describing the types of available data, ensuring that requested data is appropriate for its intended use, and limiting both the types and amount of data that researchers have access to. Data scientists 3 and 4 articulate:

> *“But as far as the research process, a researcher will usually kind of contact us, [name redacted] and I usually will kind of be, you know, between the two of us will either both of us or one of us will be the ones that kind of initiate the or have those initial conversations, to discuss kind of, you know, what are they studying? And what information do we have available that could help answer that and, and, you know, once we kind of establish that, because we are neutral, we are not data owners, we are data stewards[…]*.*”* (Data scientist 3)
>
> *“[*…*] Privacy is a big deal, you know, in this day of big data. And so, you know, we try to, we try to steer researchers to only asking for what’s absolutely necessary to do the research*.*”* (Data scientist 4)

Moreover, the workflows of data scientists centered around minimizing opportunities for bias during the application of data by researchers. During this phase of their workflow, data scientist participants expressed that working with under representative data was a challenge they faced. According to these participants, under representative data occurs as a result of missing patient identity-specific information (e.g., race and sexual orientation) and mislabeled information within patient records (e.g., gender identity). When asked why this occurs, data scientist participants explained that this was due to EHR being designed for medical billing – as opposed to disparity analysis— and EHRs limitation for capturing only those engaged in care. This challenge was similarly echoed by healthcare provider participants who also noted that EHR as a data source can only represent individuals seeking care, individuals whose records are appropriately labeled, and the individuals who have complete records. Data scientist 1 and Healthcare provider 4 explain:

> *“[…] Nobody’s looking at completeness of recording those data elements or, or accuracy or completeness or. But I do think some of the EHRs can collect that type of thing if the hospital or healthcare provider or organization that owns the data mandates that, you know, or says that that is important, and you need to record it on every patient or every client*.*”* (Data scientist 1)
>
> *“So the net that we’re getting to actually put things in the medical record is very limited by who’s actually showing up to be seen, because a lot of people don’t trust healthcare providers, or the overall system for that reason. […] I think the methods for [using EHR data] has a lot of shortcomings. Like a lot of times, [researchers and data scientists] will pull a population based on a billing code, and doctors do not always bill to match what’s clinically significant with the patient… and a lot of documentation has discrepancies between what’s clinically significant to the doctors and the way things pass through bureaucracy for processing of that person as entity within a larger financial system. So, like cause of death often does not match the cause of death, as seen by the physician. It’s usually much more vague or completely misleading, and the billing codes are often far, far short of what’s actually happening in the patient, clinically, or missed altogether*.*”* (Healthcare provider 4)

In an attempt to address this challenge, data scientists educated researchers to use external, population-level datasets to gauge how representative their data are in relation to their research question(s) of interest. Likewise, data scientists also provided researchers with education on the limitations of their requested data, education on responsible data use, guidance on data interpretation, and statistical analysis on requested datasets.

### Data Management and Utilization by Public Health Professionals

Among public health professionals, their data workflow processes were two-fold: HIV surveillance professionals’ workflows attempted to address bias through statistically completing incomplete patient records. The retention-in-care teams’ data workflows attempted to address bias through manually checking and ensuring that people living with HIV are accurately recorded as either being retained in care, or out of care.

Among HIV surveillance professionals, the receipt of incomplete and improperly collected patient records was a challenge they often experienced. Likewise, another challenge faced by these participants was receiving data that was under representative, as a result of having SDOH variables inconsistently reported by clinic staff. To address these challenges, data surveillance participants incorporated into their workflows processes for cleaning raw data, the use of statistical techniques to address issues with data quality, and trainings with field staff on appropriate data collection procedures. Health department focus group participants describe:

> *“[…]so there’s […] this constant miscommunication between the field staff and the like in our [name of health department] county clinics. Sometimes they don’t collect the data the right way, and it comes here and the [HIV surveillance] staff have to interpret some way to put it in. So there’s a lot of noise between the point of collection and the point of analysis*.*” (Health department focus group 1)*

Moreover, HIV surveillance participants noted that because their electronic reporting system is part of a larger, federal reporting system, the input fields of this reporting system are not customizable to capture local needs and did not allow interoperability with other EHR systems. This challenge led to another challenge of having data that did not representatively capture the needs of minority populations. To overcome these challenges participants emphasized understanding data limitations – in order to prevent bias in data application— and were working with other health department officials to create a new data collection system to that allows for system interoperability and the capturing local health needs. Furthermore, another challenge data surveillance professionals experienced was apprehension among community members to participate in new forms of data collection, out of fear of not knowing how state and federal governments would acquire and use their information. To overcome this, data surveillance participants reported engaging in community outreach to assuage fears and providing education to community members about new data collection methods.

Among retention-in-care specialists, the late submission of patient laboratory results was a challenge they faced. Within this workflow, this data are used to assess which individuals are retained in HIV care and which individuals required outreach to encourage them to become engaged in care. As such, when laboratory results were submitted in an untimely fashion, participants discussed being forced to make the assumption that an individual was not engaged in care, and required follow up via phone call to encourage them to return to HIV care. Health department professional 1 explains:

> *“The inability to receive timely labs [is their biggest challenge], If the physicians are not sending in their labs on time, and there’s a number of non-electronic reports that are coming in, so providers are still using the phone or paper trail to let surveillance know that a person is HIV positive and here are their labs; And they’re not automatically sending them in where it goes right into the system. [lab records] are having to be manually entered. And with that, because we [at name of engagement in care program] use surveillance systems to do our work and identify individuals, what happens is that sometimes a person may have gone to the doctor and have gotten updated labs… But based on our records those labs haven’t been entered, or they’re somewhere in a repository. And they, it causes us to call a person and say, hey, you’ve been identified as someone who is not in in medical care for a certain [mandatory reportable] condition. […]. So again, that puts us in a bad situation, and it causes heightened tension for the [person living with HIV], because they, are in their mind doing the right thing…. But our records are not up to date*.*” (Health department professional 1)*

In an attempt to overcome these challenges, retention-in-care specialists reported checking with disease surveillance professionals to check if laboratory results were submitted using alternative methods (e.g., non-electric reports). If non-electrically reported, laboratory results were then manually entered and reviewed, cross-departments, to adjust the electronic records system to better capture those retained and not-retained in care. Additionally, to overcome these challenges, retention-in-care specialists cross matched patient records with other databases, such as those within government-funded HIV clinics, state and federal justice systems, and obituaries. Health department participant 1 notes:

> *“We’re able to conduct record searches with other states to confirm if they have or have a person that mimics our demographics here in South Carolina. So, we can see if that person is in their state, and if they’re receiving care that way. We check obituaries, and we do a match with vital records to identify individuals who have been deceased. And then some people kind of link on their own, or there’re variety of ways that we go about ensuring that a person is truly not in care in South Carolina before we reach out to them…. because we don’t want to heighten their stigma, we want them to feel comfortable. We don’t want them to have some random person [contacting them] because they don’t know the staff*.*” (Health department professional 1)*

## Discussion

### Summary of Key Findings

Our study explored how workflow processes, across the data lifecycle, within HIV clinics, public health departments, and among data scientists may create or minimize opportunities for bias during EHR data collection, curation, and utilization. Throughout the stages of data collection, management, and utilization, pitfalls to minimizing EHR data bias were experienced by participants. Data collection challenges included patient-provider discomfort with soliciting and disclosing identity and sensitive health information. Data collection challenges also included difficulties and discrepancies when recording patient information in EHR systems. Moreover, during data curation and utilization, workflow challenges included receiving poor quality data and participants feeling limited in their ability to both implement and propose long-term solutions for improving data quality, data timeliness, and data standardization. Study results also showed that data workflows were structured both intentionally and unintentionally to minimize opportunities for EHR data bias. Within the context of research using EHR data, our findings are important as they illustrate EHR data’s vulnerability to social biases (e.g., structural, institutional, interpersonal), and the vulnerability of their research products to reproducing and exacerbating these biases. The implications of this are particularly important in an era where EHR data is a frequently used data source in Big Data and Artificial Intelligence research (21).

### Reducing Opportunities for Information Bias at Data Collection via Patient Portals and Satisfaction Surveys

Within the data collection stage, healthcare provider workflows were vulnerable to information bias being introduced into EHR datasets because of clinic practices and environments that were not conducive to patient information being accurately and completely recorded. For example, healthcare providers may not document patient identity and health information as a result of interpersonal and confirmation biases (22). Within our study, participants from clinical perspectives recommended increasing patient access to EHR, via patient portals, as a way to increase the accuracy and completeness of EHR data, as patients would be able to self-report and correct their demographic information. Existing literature notes improved accuracy and completeness of records when patients are given access to their EHRs (5, 13). Alpert et al. (23) in their study on oncologists’ attitudes towards patients having access to open notes within EHR systems, found that oncologists perceived this access as potentially creating a more welcoming care environment and facilitating better patient-provider communication (23). Within the context of our study population, allowing patients to enter their own information via a patient portal could help practitioners overcome discomfort with asking patients about sensitive information like demographic identity, sexual history, and sexual practices. Additionally, the possibility of improved EHR accuracy and completeness could mean an improved ability to identify and address proximal and distal causes to health outcomes stemming from structural biases (24, 25). Therefore, future research is needed to understand possible differences in sensitivity when using patient-portal accessible EHR records, versus non-patient portal accessible records, to identify potential structural biases in health outcomes.

Despite its potential, it is important to understand possible barriers to successfully implementing patient portals as way to improve data accuracy and completeness. Gybel Jensen et al. (26), in their study on patients’ experiences with digitalization in a health system, found that when e-Health interventions (e.g., patient portals to their EHRs) were not tailored to meet the needs, health literacy, and tech literacy of patients, they were perceived by patients as being inaccessible (26). Choy et al. (27), in their study on the digital health experiences of patients in primary care, found that successful e-Health interventions (e.g., patient EHR portals) needed to be implemented with an understanding of how to overcome barriers to internet access often faced by individuals with a low socioeconomic status (27). Therefore, further implementation science studies are warranted to establish equitable access to patient portals to increase the accuracy and completeness of EHR data for patients of all backgrounds.

In addition to accessible patient portals, study participants from healthcare perspectives also suggested the use patent-care-experience surveys as a tool to improve EHR data completeness and accuracy. Specifically, these participants perceived patient-care-experience surveys as a way to help healthcare professionals and health system administrators identify and address dynamics impacting patient comfort with disclosing sensitive health behavior information (e.g., healthcare provider treatment, timing and sequences of clinical processes). Extant literature observes the ability of patient experience surveys to effectuate change as being mixed, with cost efficiency documented as a notable challenge (28). Despite these challenges, patient experience surveys are perceived by the existing literature as being associated with better clinical outcomes and patient safety (29). The literature also notes that successful patient experience surveys must be developed with the clear intention of meeting a specific goal (e.g., understanding and reducing experiences of biased healthcare provision) and tailored to suit the local contexts where it is being deployed (28). As such, we posit that patient-care-experience surveys, when developed and implemented according to best practices, could be a viable tool, both in the immediacy and the long-term, to identify and address interpersonal, institutional, and structural biases occurring within HIV-specific care settings, subsequently warranting future research.

### Reducing Underrepresentation in EHR Data Through Improving Access and Engagement in Care

Data scientists and health department professionals in our study noted underrepresentation as a challenge they faced when using EHR data to draw statistical conclusions and developing public health surveillance-based decision-making tools. These participants perceived underrepresentation as occurring as a result of incomplete demographic information and because of EHR’s ability to only capture individuals engaged in care. Existing literature offers statistical techniques, using external datasets, to assess the representativeness of EHR data (10-13). However, our study results, situated within a deep south context, suggest that it is also important to proactively address underrepresentation through increasing the affordability, availability, and accessibility of care for populations not engaged in care (30). As such, we recommend that further research is needed on innovative strategies to engage stigmatized and marginalized populations that have been historically hard to engage in HIV care (e.g., undocumented individuals, young men who have sex with men, individuals engaging in illicit substance use).

Studies within the health contexts of cardiovascular disease, maternal health, and obesity note that patients represented in EHR systems are more likely to be ill, leading to informed presence bias, and therefore complicating the generalizability of EHR-derived results to the broader population (13, 31, 32). As HIV is a mandatory reportable condition in the United States, except in Idaho and the U.S. Virgin Islands, its diagnosis and the progress towards its management must be reported by all healthcare practitioners, to state health departments, regardless of an individuals’ health status (33). Therefore, HIV studies using EHR data may be more generalizable at a population level, than other disease contexts. Our findings suggest that informed presence bias, a form of selection bias, may not be operating within HIV-specific EHR datasets to the same extent that it would within other chronic disease contexts. However, not all people living with HIV are linked to HIV care services in a timely manner. As such, because there is a high likelihood that the most vulnerable living with HIV (e.g., unhoused populations, people who use illicit substances) face limited access to healthcare due to stigma, lack of health insurance, and other adversities, their information most likely would not be included in existing EHR systems or subsequent datasets. As such, future research is needed on the representation of vulnerable populations living with HIV and the generalizability of HIV-specific EHR data at state, regional, and national levels.

### Increasing Opportunities for Stakeholder Collaboration to Reduce Opportunities for EHR Data Bias

Participants whose workflows occurred during data curation and utilization phases described untimely data submission, the non-uniform submission of data, and the submission of inaccurate and incomplete records as instances where bias was introduced; and as challenges they faced when minimizing opportunities for statistical bias. To improve data quality, health department participants described the need to communicate with healthcare providers about standards for reporting HIV diagnoses and patient progress towards viral suppression (e.g., timeliness, reporting format, completeness of data, types of information requested). This is in addition to their efforts to provide training to increase healthcare provider knowledge of these standards. However, these participants noted data quality as an ongoing issue and were limited in how much change they could affect because cross-facility and cross-department protocols fell outside the scope of their roles. Furthermore, our study found that clinics providing care solely to people living with HIV (e.g., Ryan White) were anecdotally perceived as being more adherent to HIV reporting standards, in comparison to private healthcare providers. These findings reflect that of Sauer et al. (10) and Charpignon et al. (34) who assert that to improve the quality of EHR data, we need to understand the best practices for strengthening working relationships between clinicians and data scientists (10, 34). In addition, further research is needed to understand how HIV reporting standard adherence may differ between Ryan White-based healthcare providers and private healthcare providers. Similarly, it is important to understand how challenges and incentives to meet HIV-specific reporting standards may differ within these two healthcare contexts.

### Methodological Considerations

This study has both strengths and limitations. Transferability was increased through providing thick descriptions of participant perspectives via transcript quotations (35). Our study results may be applicable to other settings in the South Eastern United States that similarly resemble South Carolina regarding its HIV disease burden, infectious disease reporting practices, and health system structure. Our study results may also be applicable to similar settings in the Southeast using EHR systems with limited integration and limited practices in place to address social and statistical biases. Furthermore, confirmability was increased through using a reflexive journal, and the use of an analysis matrix (35). Credibility was increased through the use of an audit trail (35). Lastly, confirmability, credibility, and dependability were increased through triangulation via collecting data from multiple perspectives (i.e., healthcare providers, patients, data scientists, and public health professionals) (35). A limitation of this study is that saturation was not reached (16). As a result, the inclusion of additional stakeholder and participant perspectives (e.g., public health researchers) could have led to additional findings that are not captured within this study. As such, we recommend that future studies on efforts to reduce EHR data bias include the perspectives of researchers using the EHR datasets provided by data scientists.

## Conclusion

Regardless of how data collection, curation, and utilization workflow processes were structured, challenges and pitfalls within each workflow created opportunities for EHR data bias to occur. The study results found that opportunities for bias identified by participants stemmed from the number of times EHR data changed hands (e.g., different healthcare providers entering patient information; data scientists and public health surveillance professionals cleaning and analyzing data; researchers and public health professionals creating products and making decisions from analyzed data) and the limited oversight each stakeholder perspective had on others and their workflows when engaging with EHR data. To our knowledge, this study is the first to explore how workflow processes, within and between HIV clinics, public health departments, and data scientists, may identify and address opportunities for bias during EHR data collection, curation, and utilization (Gagnon et al., 2023; Kelly et al., 2023; Skolnik, 2020). Within a larger public health context our results can be used to inform healthcare system and public health policies focused on improving the quality of EHR data. Likewise, the results of our study can also be used to aid stakeholders in their informed decision making on both how and the degree to which they use EHR data in their research and public health products.

## Data Availability

All data produced in the present study are available upon reasonable request to the authors.

## Declarations

## Acknowledgements

We would like to thank the participants in our study for sharing their experiences with us. We would also like to thank Audrey Auen and Miranda Nixon for their help with interviewing study participants. Lastly, we would like to thank Ms. Christina Miller for her help coordinating participant interviews.

## Author Approval

All authors have reviewed and approved the manuscript.

## Ethics

This study received IRB approval from The University of South Carolina Office of Research Compliance (Application No.: Pro00122501) and was conducted in accordance with the Declaration of Helsinki.

## Data Availability

All data produced in the present study are available upon reasonable request to the authors.

